# Psychotic experiences and disorders in adolescents and young adults with intellectual disabilities: Evidence from a population-based birth cohort in the United Kingdom

**DOI:** 10.1101/2024.07.10.24310201

**Authors:** Christina Dardani, Jack F G Underwood, Hannah J. Jones, Alexandros Rammos, Sarah Sullivan, Laura Hull, Golam M. Khandaker, Stan Zammit, Dheeraj Rai, Paul Madley-Dowd

**Affiliations:** MRC Integrative Epidemiology Unit, Bristol Medical School, University of Bristol, UK; Population Health Sciences, Bristol Medical School, University of Bristol, UK; Division of Psychological Medicine and Clinical Neurosciences, School of Medicine, Cardiff University, UK; National Institute for Health Research Biomedical Research Centre, University of Bristol, UK; Department of Clinical, Educational & Health Psychology, University College London, London, UK; Bristol Autism Spectrum Service, Avon and Wiltshire Partnership NHS Mental Health Trust, Bristol, UK

## Abstract

**Background:** Evidence suggests that individuals with intellectual disabilities may be at increased risk of affective and non-affective psychotic disorders. However, studies so far have been limited by small and selected samples. Moreover, the role of early life trauma, a key environmental risk factor for psychosis, in the potential associations is unknown.

**Methods:** Using data from Avon Longitudinal Study of Parents and Children (ALSPAC) birth cohort, we investigated the associations between ID, psychotic disorders, and psychotic experiences in adulthood, and assessed the potential mediating role of trauma in childhood. Individuals with intellectual disabilities were identified using a multisource measure utilising cognitive, functional, and diagnostic indicators from ALSPAC combined with health and administrative records. Psychotic disorder clinical diagnoses were extracted through linkage to primary care records. Psychotic experiences were assessed at ages 18 and 24 using the semi-structured Psychosis-Like Symptoms interview (PLIKSi). Traumatic experiences between ages 5 and 11 were assessed with questionnaires and interviews administered to children and parents at multiple ages. Multiple imputation was performed across all analyses to mitigate potential bias due to missing data.

**Findings:** The maximum sample after multiple imputation was 9,407. We found evidence of associations between intellectual disabilities and psychotic disorders (OR= 4.57; 95%CI: 1.56-13.39). Evidence was weaker in the case of psychotic experiences (OR=1.63; 95%CI: 0.93-2.84). There was some evidence suggesting a potential mediating role of traumatic experiences in the associations between ID and psychotic experiences (OR= 1.09; 95%CI: 1.03-1.15). Evidence was less consistent in the case of psychotic disorders. Complete records analyses yielded comparable estimates.

**Interpretation:** Intellectual disabilities are associated with psychotic disorders and experiences in young adulthood. Further research into the contribution of trauma could shape current intervention strategies for psychotic disorders in this population.

**Funding:** The Baily Thomas Charitable Fund

## Introduction

Intellectual Disability (ID) refers to difficulties in cognitive and adaptive functioning that manifest early in childhood and have a substantial impact on education, independent living skills, employment, and access to social support and health care across the lifespan^1^. ID has a neurodevelopmental origin and is not the result of later neurocognitive changes due to injury or disease. Recent meta-analytic evidence suggests that lifetime prevalence of mental health conditions in ID may be substantially higher than the general population (pooled lifetime prevalence: ≈32% in individuals with ID vs ≈29% in the general population)^2,3^. Co-occurring mental health conditions in ID have been associated with adverse behavioural, educational, and social outcomes for the affected individuals as well as lower quality of life for their families and carers^4^. On this basis, understanding the links between ID and mental health conditions is among the top global research priorities in the field^5^.

Affective and non-affective psychotic disorders (henceforth psychotic disorders) are among the most common co-occurring mental health conditions in ID. The prevalence of psychotic disorders in ID, appears to be higher than their estimated lifetime prevalence in the general population, with schizophrenia reaching approximately 4.8% (vs ≈0.9% in the general population), unspecified psychotic disorder reaching 3.9% (vs ≈0.5%) and bipolar disorder approximately 2% (vs ≈0.2%)^2,6–8^. However, most studies so far have been limited by small and selected samples (predominantly inpatient), with limited control for potential confounding factors. Furthermore, these studies have been predominantly based on diagnoses of psychotic disorders in this population^2^, offering limited insights into sub-clinical symptoms and potential precursors of psychotic disorders, such as psychotic experiences.

Several factors have been proposed to influence risk of psychiatric conditions in individuals with ID, including access to support services, poor physical health, and major life events (including but not limited to trauma)^9,10^. The latter is particularly relevant in the case of psychotic disorders. Traumatic life events (such as physical, sexual and emotional abuse, neglect, domestic violence and bullying victimisation) are among the most consistently identified risk factors for psychotic experiences and disorders in the general population^11,12^. Emerging evidence suggests high rates of exposure to traumatic life events in the ID population^13,14^. There is an absence of studies investigating whether and to what extent traumatic life events mediate any associations between ID and psychotic experiences and disorders.

Using prospectively collected questionnaire, interview and health record linkage data in a population-based birth cohort in the UK, we assessed: (A) the risk of psychotic disorders and psychotic experiences during early adulthood in individuals with and without ID, (B) the potential associations of ID to longitudinal profiles of psychotic experiences reflecting the persistence and frequency of psychotic experiences from age 12 to age 24 (C) the extent to which trauma experienced in childhood may mediate the links between ID and psychotic disorders and psychotic experiences, (D) the possible confounding influence of familial, socioeconomic and demographic factors in any identified links.

**Research in context**

**Evidence before this study**

Evidence suggests that individuals with Intellectual Disability (ID) may be at increased risk of affective and non-affective psychotic disorders (henceforth psychotic disorders).

We searched PubMed for studies published in English before March 11, 2024, using the terms (“psychosis” OR “psychotic” OR “bipolar” OR “mani*” OR “schiz*”) AND (“intellectual disabilit*”). Among the studies that our search strategy yielded, we identified three meta-analyses aiming to estimate the prevalence of mental health conditions, including affective and non-affective psychotic disorders, in individuals with ID.

All three meta-analyses indicated a higher than general population prevalence of affective and non-affective psychotic disorders in ID. However, most of the studies included in the meta-analyses had been limited by small and selected samples (predominantly inpatient), with minimal control over potential confounding factors. This is in contrast to research into psychotic disorders in the general population, where investigations have expanded to assess links to sub-clinical manifestations of psychotic disorders, psychotic experiences, and have identified a number of potential factors mediating risk such as trauma in childhood.

**Added value of this study**

In the present study we adopted the research paradigms of studies investigating risk of psychotic disorders in the general population and interrogated whether ID is associated with psychotic disorders and psychotic experiences. We additionally investigated the potential mediating role of traumatic experiences in childhood. We used prospectively collected questionnaire, interview, and health record linkage data in a population-based birth cohort. Our findings suggest that ID may be associated with psychotic disorders and experiences in young adulthood and trauma may at least partially mediate some of the associations.

**Implications of all the available evidence**

Our findings encourage further research into the factors influencing risk of psychotic disorders and experiences in individuals with ID, particularly with regards to trauma. This line of research is expected to shape current intervention approaches for psychotic disorders in this the ID population by stimulating a shift towards trauma-focused therapies.

## Methods

### Study Design and Participants

A visual summary of the study aims, and design can be found in Figure 1. The Avon Longitudinal Study of Parents and Children (ALSPAC) is a population-based cohort study of children born to 14,541 pregnant mothers residing in the former county of Avon, United Kingdom, with an expected delivery date between 1 April 1991 and 31 December 1992. Of these initial pregnancies, there was a total of 14,676 foetuses, resulting in 14,062 live births and 13,988 children who were alive at 1 year of age. When the oldest children were approximately 7 years of age, eligible samples who did not join the study initially were contacted, and additional participants were recruited. This resulted in a total of 15,447 pregnancies and 15,658 foetuses, of which 14,901 were alive at 1 year of age.

**Figure 1.**
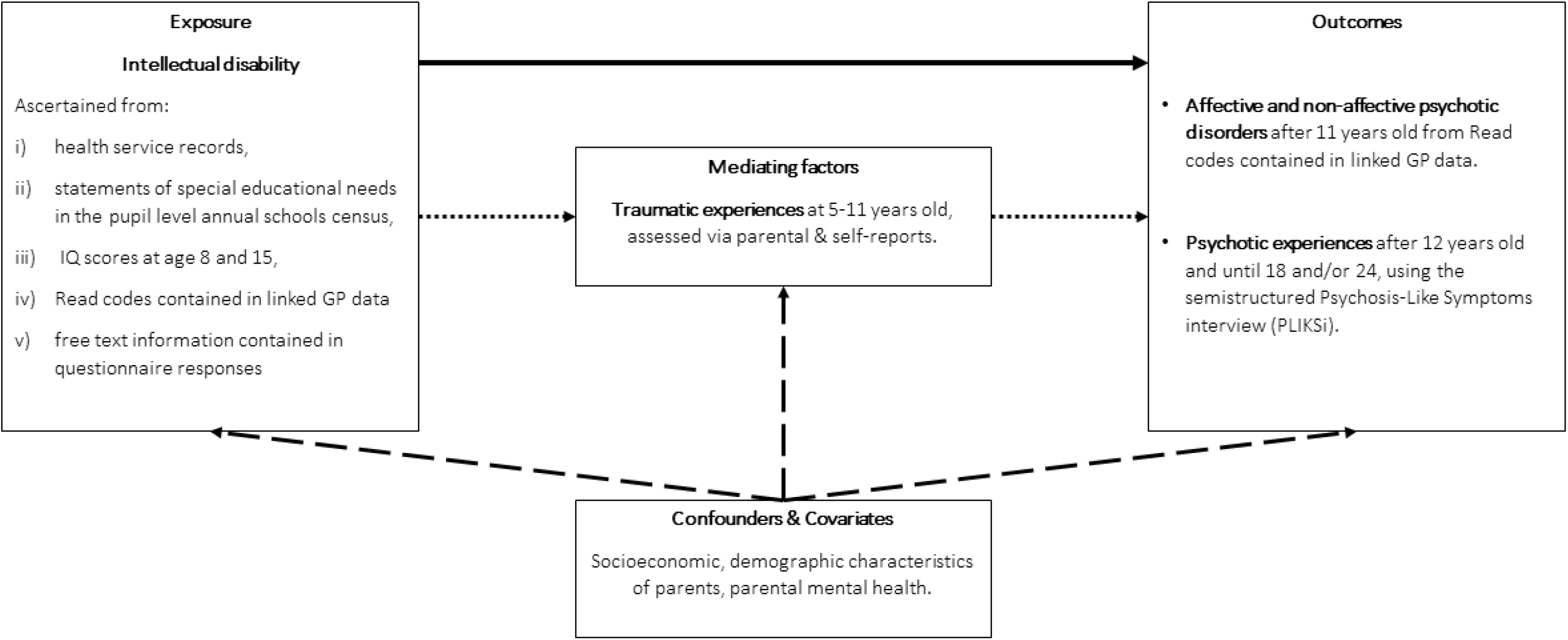
Visual summary of the study aims and design. Solid black lines correspond to the analyses investigating the links between intellectual disabilities (ID) and psychotic disorders and experiences. Dotted black lines correspond to the analyses investigating the extent to which any links between ID and psychotic disorders and experiences are mediated via traumatic experiences in childhood. Dashed black lines correspond to the analyses investigating the possible confounding influence of familial, socioeconomic, and demographic factors in the links between ID and psychotic disorders and experiences. Please note that ID is typically of neurodevelopmental origin and therefore in the present study it was assumed to precede the mediator and outcome.

Further information on the ALSPAC cohort is available on the ALSPAC website (http://www.bristol.ac.uk/alspac) and elsewhere.^15–17^ The study website contains details of all the data that is available through a fully searchable data dictionary and variable search tool (http://www.bristol.ac.uk/alspac/researchers/our-data/). Some study data were collected and managed using REDCap electronic data capture tools hosted at the University of Bristol. REDCap (Research Electronic Data Capture) is a secure, web-based software platform designed to support data capture for research studies.^18,19^

For data collected via questionnaires and clinics, informed consent was obtained from participants following the recommendations of the ALSPAC Ethics and Law Committee at the time. Ethical approval for the study was obtained from the ALSPAC Ethics and Law Committee and the Local Research Ethics Committees (NHS Haydock REC: 10/H1010/70).

### Linkage to health and administrative records

When the index children reached legal adulthood (age 18), ALSPAC conducted a postal fair-processing campaign to re-enrol them into the study and to seek permission for linkage to health and administrative records. This was an ‘opt out’ approach, meaning linkage was attempted for all participants, except those who objected and those who were not sent fair processing materials. Where ‘opt in’ consent became practicable (e.g., when a participant attended a study assessment visit) then this was collected by a trained fieldworker. Details on the linkage process can be found elsewhere^20^.

In the context of the present study, the eligible sample was defined based on the following criteria: 1. having available linkage data and not dissenting to their use, 2. having linked primary care data available from age 11 onwards (considering that we were interested in the adolescent and early adulthood period). A total of 9,680 participants were eligible.

## Measures

### Exposure: ID

ID was identified in ALSPAC using a composite measure created in previous published work^21^ based on information from six different sources: i) IQ scores less than 85 measured at age 8 and at age 15, ii) free text fields from parent reported questionnaires, iii) school reported provision of educational services for individuals with a statement of special educational needs for cognitive impairments, iv) from relevant Read codes^22^ contained in GP records (Read codes are a comprehensive list of standardised clinical terms used by healthcare professionals within the UK National Health Service to record clinical information), v) international classification of disease (ICD) diagnoses contained in hospital episode statistics and vi) recorded interactions with mental health services for ID contained within the mental health services data set. The ID measure is available on the UK Secure eResearch Platform (UKSeRP). Further details on the creation of the composite ID measure can be found in the original publication^21^.

### Primary Outcome: Psychotic disorders

Read codes (V.2) from GP records relevant to diagnosis and symptoms of psychotic disorders were extracted to identify the outcomes of interest in the eligible sample. GP records were available from 1990 to 2016, when the oldest participants were 25 years of age. The full list of Read codes used can be found in the project dedicated repository: https://github.com/pmadleydowd/BailyThomas-IntellectualDisability-and-MentalHealth.

### Primary Outcome: Psychotic experiences until early adulthood

Psychotic experiences were assessed at ages 18 and 24 using the semi-semi structured Psychosis-Like Symptoms interview (PLIKSi), administered by trained psychologists, and scored according to criteria predefined by the World Health Organization^23^. The PLIKSi consists of 12 core questions covering hallucinations, delusions, and thought interference. Participants were asked about experiences that had occurred since age 12 years. Psychotic experiences were considered present if, at ages 18 and/or 24 years, one or more of the experiences was rated by the interviewer as suspected or definitely present, and if this was not attributable to falling asleep or waking up or fever. We additionally examined psychotic experiences that had been distressing and/or frequent, since these experiences are more clinically relevant and predictive of psychotic disorder^24^.

### Secondary Outcome: Longitudinal profiles of psychotic experiences

Considering that psychotic experiences are, in most cases, transient in the general population^24^, and do not necessarily reflect liability to psychotic disorders later in life, we additionally used a measure reflecting the persistence and frequency of psychotic experiences across three time points in ALSPAC: ages 12, 18 and 24 years. Details on the measure can be found in the original publication^25^. Briefly, using information from the PLIKSi on current presence (over past 6 months) and frequency of psychotic experiences (0: “Not present”, 1: “Low-frequency”-experiences occurring less than weekly, 2: “High-frequency”-experiences occurring weekly or daily), at each time point, an empirical composite measure was generated reflecting four longitudinal profiles of psychotic experiences from ages 12 to 24: A. No experiences: Individuals without a psychotic experience at any time point; B. Transient: Individuals with a psychotic experience rated at only one time-point, regardless of frequency; C. Low-frequency persistent: Individuals with low-frequency psychotic experiences at two or more time points, or with a low-frequency rating at one time point and a high-frequency rating at another; D. High-frequency persistent: Individuals with high-frequency psychotic experiences rated at two or more time points. Following previous work we did not make an assumption on the potential severity ordering of the profiles^25^, particularly considering that the boundaries between the transient and persistent-low profiles might be difficult to define.

### Mediator: Traumatic experiences in childhood

The measures of childhood trauma and their associations with psychotic experiences have been described in detail elsewhere^11^. In brief, we used a measure of childhood trauma between ages 5 and 11 based on responses to 57 questions from questionnaires and interviews about domestic violence (regular acts of physical violence taking place in the home), physical abuse (physical harm to the participant from caregivers or other adults), emotional abuse (emotional cruelty to the participant from caregivers or other adults), emotional neglect (caregivers not taking an interest in the participant’s life), sexual abuse (adults or older children forcing the participant into sexual activity, including attempts to do so), and bullying victimization (regular name-calling, blackmail, or assault by peers). Measures of sexual, physical, and emotional abuse, assessed contemporaneously by the participant and their caregivers between participant ages 5 to 11, were supplemented with data from a participant-completed questionnaire at age 22, as all data on sexual abuse, and most data on physical and emotional abuse prior to age 11, were based on parental report. Each type of trauma was coded as present or not, and a single trauma variable was created representing exposure to any type of trauma^11^.

### Covariates

Covariates in the present study were selected on the basis of their potential associations with the exposure, outcomes and mediator. These included child sex (male/female), maternal parity (≤1 child versus ≥ 2 children), major financial problems in the family when the child was 8 months old (yes/no), maternal highest educational attainment (32 weeks gestation), maternal age (at delivery), maternal Crown-Crisp anxiety scores^26^ (18 weeks gestation), and maternal depression measured with the Edinburgh Postnatal Depression Scale^27^ (EPDS; 18 weeks gestation scores ≥ 13).

### Statistical Analyses Association analyses

Statistical analyses were conducted in StataSE version 18. We estimated descriptive statistics of participant characteristics for individuals with and without ID, traumatic experiences, and psychotic experiences.

Using logistic regression, we estimated odds ratios (OR) and 95% confidence intervals (95% CI) for the associations between ID and psychotic disorders as well as psychotic experiences in early adulthood. Using g-computation via Stata’s margins command, we further estimated the adjusted marginal risk (over all covariates) and risk difference of each outcome for participants with and without ID. Standard errors were calculated using the delta method. Using multinomial logistic regression, we estimated relative risk ratios (RRRs) and 95% CIs for the associations between ID and the four longitudinal profiles of psychotic experiences. Across all association analyses we performed crude and covariate-adjusted models.

### Mediation analyses

We decided *a priori* to conduct mediation analyses between ID (exposure), trauma (mediator) and psychosis related outcomes regardless of whether there was evidence of associations between ID and the outcomes of interest. This decision was based on previous work suggesting that evidence of associations between exposure and outcome should not guide decisions for subsequent mediation analyses, particularly when the effect size is expected to be small or there may be suppression effects (when the direct and indirect effects of an exposure on an outcome have opposite directions)^28,29^. Mediation analyses were performed using the g-formula package^30^ in Stata. We used the parametric g-formula using 10,000 Monte Carlo simulations to estimate the natural direct effect (NDE) of ID on psychotic experiences, and the natural indirect effect (NIE) that was mediated via traumatic experiences between ages 5 and 11. We performed crude as well as covariate-adjusted models. Corresponding 95% CIs were estimated using the standard errors from 1000 nonparametric bootstrap resamples.

### Missing data

Considering previous work suggesting that individuals with ID are more likely to have missing data in ALSPAC (particularly those with more severe ID)^21^, we decided *a priori* to perform multiple imputation across all association and mediation analyses to mitigate potential bias from missing data ^31^.For primary and secondary outcomes, we performed multiple imputation by chained equations, using Stata’s *MI impute* command. One hundred datasets were imputed with 25 burn-in iterations and estimates were combined across imputed datasets using Rubin’s rules, implemented via Stata’s *MI estimate* command. We included auxiliary variables to make the missing at random assumption more plausible^32^. Based on established guidelines on auxiliary variables selection we entered in the models those variables presenting the lowest missingness in the eligible sample, ranging from 12-13%. Details on the imputation models applied and the auxiliary variables used can be found in Supplementary Note 1. In the case of mediation analyses, we used the inbuilt g-formula imputation commands allowing simultaneous imputation of missing data and mediation analyses, entering in the models the same auxiliary variables we used for the association analyses. In the context of the present study, we present both complete records and imputed data analyses, although we consider as primary the imputed data analyses.

## Results

The maximum sample size with data on exposure and at least one outcome measure was 9,407 (49.6% male; 3.6% ID; 0.3% psychotic disorder diagnosis). Full characteristics of our study sample, including auxiliary and outcome variables are listed in Supplementary Table 1. Those with ID were more likely to have experienced trauma between ages 5-11. The mothers of those with ID were less likely to have a university degree and had a greater prevalence of screening positive for depression. Approximately 66% of the sample had complete data on exposure, psychotic disorder diagnosis (complete for all participants in the eligible sample), and covariates, while 33% of the sample had complete data on exposure, psychotic experiences, and covariates. Participants with complete data were more likely to have a higher socioeconomic background than those with incomplete (details on the identified patterns can be found in Supplementary Tables 2a & 2b).

### Association analyses

There was some evidence suggesting an association between ID and primary care diagnoses of psychotic disorders in crude and adjusted for covariate models (adjusted OR=4.57; 95%CI: 1.56, 13.39; Table 1). When considered on the risk difference scale, this odds ratio reflects a small absolute increase in risk among those with ID (adjusted marginal risk difference= 1.07%; 95%CI:-0.29%, 2.44%; Table 1); we were unable to report the absolute risk according to ID group due to low counts with a diagnosis among individuals with an ID. There was also some evidence to support associations between ID and psychotic experiences (adjusted OR=1.63; 95%CI: 0.93, 2.84; adjusted marginal risk difference=8.41%; 95%CI:-2.21%, 19.03%; Table 1) as well as distressing and/or frequent psychotic experiences (adjusted OR=1.92; 95%CI: 0.96, 3.86; adjusted marginal risk difference=8.88%; 95%CI:-2.44%, 20.20%; Table 1) although the confidence intervals of these results crossed the null. Association estimates in complete records analyses were of comparable magnitude, albeit less precise (Supplementary Table 3).

**Table 1.**
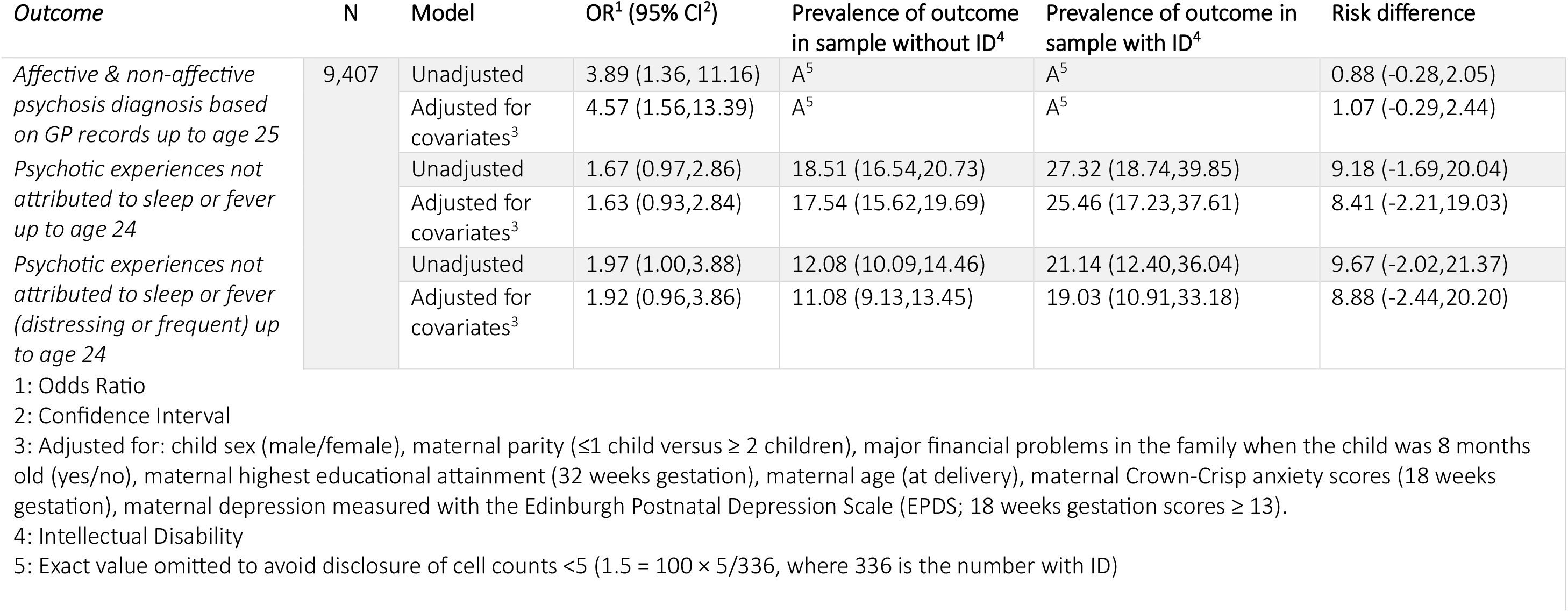
Associations between intellectual disability, psychotic disorders, and psychotic experiences in early adulthood from multiple imputation analyses.

There was little evidence to suggest that individuals with ID may be more likely to present with persistent profiles of psychotic experiences. Although the relative risk ratios for the high frequency persistent profiles were larger than those for the low frequency persistent and transient profiles, the estimates were highly imprecise (Table 2 for imputed data analyses and Supplementary Table 4 for complete records analyses).

**Table 2.** Associations between ID and longitudinal profiles of psychotic experiences from multiple imputation analyses.

| *Outcome* | *N* | *Model* | *RRR<sup>1</sup> (95% CI<sup>2</sup>)* |
| --- | --- | --- | --- |
| *No psychotic experiences* | 9,407 | Unadjusted | Ref |
| *Persistent High* |  |  | 3.14 (0.65,15.06) |
| *Persistent Low* |  |  | 1.21 (0.45,3.21) |
| *Transient* |  |  | 1.55 (0.85,2.84) |
| *No psychotic experiences* |  | Adjusted for covariates<sup>3</sup> | Ref |
| *Persistent High* |  |  | 2.85 (0.54,15.01) |
| *Persistent Low* |  |  | 1.20 (0.44,3.27) |
| *Transient* |  |  | 1.49 (0.80,2.79) |

1: Relative Risk Ratio
2: Confidence Interval
3: Adjusted for: child sex (male/female), maternal parity ( $\leq 1$ child versus $\geq 2$ children), major financial problems in the family when the child was 8 months old (yes/no), maternal highest educational attainment (32 weeks gestation), maternal age (at delivery), maternal Crown-Crisp anxiety scores (18 weeks gestation), maternal depression measured with the Edinburgh Postnatal Depression Scale (EPDS; 18 weeks gestation scores $\geq 13$ ).

### Mediation analyses

There was some evidence to suggest that childhood trauma may mediate the associations between ID and psychotic experiences (effect of exposure on outcome via the mediator, NIE, adjusted OR= 1.09; 95%CI: 1.03-1.15), as well as distressing and/or frequent psychotic experiences (effect of exposure on outcome via the mediator, NIE, adjusted OR= 1.11; 95%CI: 1.03-1.20). Evidence was weaker in the case of psychotic disorders, where traumatic experiences did not appear to mediate the associations with ID (Table 3 for imputed data analyses and Supplementary Table 5 for complete records analyses).

**Table 3.** Results of the mediation analyses with childhood traumatic experiences, for the associations between ID, psychotic disorders and psychotic experiences using imputed data.

| *Outcome* | *Model* | *N* | *TCE<sup>1</sup>; OR<sup>2</sup> (95%CI<sup>3</sup>)* | *NDE<sup>4</sup>; OR<sup>2</sup> (95%CI<sup>3</sup>)* | *NIE<sup>5</sup>; OR<sup>2</sup> (95%CI<sup>3</sup>)* |
| --- | --- | --- | --- | --- | --- |
| *Affective & non-affective psychosis diagnosis based on GP records up to age 25* | Crude | 9,407 | 3.90 (1.28, 11.90) | 3.64 (1.19, 11.11) | 1.07 (0.98, 1.17) |
|  | Adjusted<sup>6</sup> |  | 5.82 (1.83, 18.53) | 5.53 (1.71, 17.91) | 1.03 (0.85, 1.26) |
| *Psychotic experiences not attributed to sleep or fever up to age 24* | Crude |  | 0.95 (0.62, 1.46) | 0.89 (0.58, 1.36) | 1.07 (1.01, 1.13) |
|  | Adjusted<sup>6</sup> |  | 1.07 (0.64, 1.78) | 0.98 (0.59, 1.63) | 1.09 (1.03, 1.15) |
| *Psychotic experiences not attributed to sleep or fever (distressing or frequent) up to age 24* | Crude |  | 0.99 (0.62, 1.59) | 0.91 (0.57, 1.45) | 1.09 (1.01, 1.17) |
|  | Adjusted<sup>6</sup> |  | 1.19 (0.61, 2.34) | 1.07 (0.54, 2.11) | 1.11 (1.03, 1.20) |

1: Total effect
2: Odds Ratio
3: Confidence Intervals
4: Natural Direct Effect
5: Natural Indirect Effect
6: Adjusted for: child sex (male/female), maternal parity ( $\leq 1$ child versus $\geq 2$ children), major financial problems in the family when the child was 8 months old (yes/no), maternal highest educational attainment (32 weeks gestation), maternal age (at delivery), maternal Crown-Crisp anxiety scores (18 weeks gestation), maternal depression measured with the Edinburgh Postnatal Depression Scale (EPDS; 18 weeks gestation scores $\geq 13$ ).

## Discussion

Using prospectively collected questionnaire, interview, and health record linkage data in a population-based birth cohort, we examined the associations between ID, psychotic disorders, and psychotic experiences in early adulthood and investigated the factors that may influence them. We found evidence suggesting that ID may be associated with psychotic disorders. Although evidence was less consistent in the case of psychotic experiences, traumatic experiences in childhood appeared to mediate their associations with ID. The identified relationships were unlikely to be explained by familial, socioeconomic, and demographic factors.

### The relationships of ID to psychotic disorders and experiences

Our findings are consistent with a growing body of evidence suggesting that people with ID may be at higher risk of psychotic disorders than the general population. The latest and largest study in the field (N=2,091) found that individuals with ID had higher risk of psychosis not only compared to the general population, but also compared to individuals with other neurodevelopmental conditions (e.g., autism, ADHD)^33^. There is an ongoing discussion on the possibility of bias in the existing evidence due to confounding and/or measurement error (measurement error might arise for example, due to the application of diagnostic criteria and tools designed for the general population in individuals with ID)^8^. In our study we attempted to overcome confounding bias by adjusting our models for several familial, socioeconomic, and demographic factors and found that they are unlikely to explain the identified links (although the possibility of residual confounding cannot be excluded-see Strengths and limitations section). With regards to measurement error, evidence of associations with psychotic disorder diagnoses were complemented with some evidence of associations between ID and subclinical expressions of psychosis liability, psychotic experiences. Although the evidence was relatively inconsistent across psychotic experiences measures, the direction of the association estimates was consistent with the ones identified for psychotic disorders. Moreover, the strongest associations were identified in the case of psychotic experiences that were distressing and/or frequent, a phenotype that is considered to be more strongly related to subsequent risk of psychotic disorders^24^.

### The mediating role of traumatic experiences in childhood

Our study provides some evidence on the potentially mediating role of traumatic experiences in childhood in the associations between ID and psychotic experiences. It is worth noting here that evidence was weaker in the case of psychotic disorder possibly due to the lack of power, particularly considering that only 0.3% of the total eligible sample (36 individuals) had a diagnosis of psychotic disorder.

Although our study is the first to apply a formal counterfactual mediation approach, previous work in a sample of 1,023 adults with ID found that major life events (including but not limited to trauma) were associated with psychiatric conditions in this population^10^. On this basis, interventions for trauma-related morbidity in this population may substantially improve mental health outcomes. Evidence on the effectiveness of trauma-focused interventions in people with ID is promising, indicating that eye movement desensitization and reprocessing (EMDR) as well as trauma-focused cognitive behavioural therapy (CBT) may be effective in this population^34^, with substantive trials underway (e.g., https://www.isrctn.com/ISRCTN35167485). However, most of the evidence so far comes from case-studies and therefore further work is necessary to appraise the appropriateness and effectiveness of these interventions in people with ID.

### Strengths and limitations

This is the first study to investigate the links between ID, psychotic disorders and psychotic experiences and assess the possible influence of traumatic experiences in childhood using prospectively collected data from a large population-based cohort. We also used linkage to health and administrative record data, which aided the identification of ID and psychotic disorder cases and reduced the impact of attrition and therefore bias due to missing data.

Our study presents several limitations. First, our exposure definition includes both mild and more severe cases of intellectual disability and associations may differ by the severity of ID. Second, although we used psychotic experiences as a phenotype reflecting psychotic disorder liability, psychotic experiences are associated with several adverse mental health outcomes such as depression^35,36^. Third, our complete records analyses may have been limited by lack of power. Fourth, although we tried to mitigate the possibility of bias due to missing data using multiple imputation, some bias is still likely to influence the findings of the analyses using psychotic experiences as the outcome (association & and mediation analyses). This is because individuals with ID and psychosis are less likely to participate in ALSPAC and the use of psychotic disorder diagnoses as an auxiliary for psychotic experience is unlikely to fully break the link between the outcome and the probability of missing data^37^. Fifth, although we adjusted our analyses for a number of potential familial, socioeconomic, and demographic factors, some level of residual confounding is still likely to be present.

## Conclusions

ID is associated with psychotic disorders and experiences into young adulthood a. Traumatic experiences in childhood may partially mediate the associations and further research in this area could shape current intervention strategies for psychotic disorders in this population.

## Data Availability

Individual-level data from the ALSPAC birth cohort are not publicly available for reasons of clinical confidentiality. Data can be accessed after application to the ALSPAC Executive Team who will respond within 10 working days. Application instructions and data use agreements are available at http://www.bristol.ac.uk/alspac/researchers/access/. The code used to conduct these analyses can be found in the repository: https://github.com/pmadleydowd/BailyThomas-IntellectualDisability-and-MentalHealth

## Acknowledgements

We are extremely grateful to all the families who took part in this study, the midwives for their help in recruiting them, and the whole ALSPAC team, which includes interviewers, computer and laboratory technicians, clerical workers, research scientists, volunteers, managers, receptionists, and nurses. The UK Medical Research Council and Wellcome (Grant ref: 217065/Z/19/Z) and the University of Bristol provide core support for ALSPAC. A comprehensive list of grants funding is available on the ALSPAC website (http://www.bristol.ac.uk/alspac/external/documents/grant-acknowledgements.pdf). This publication is the work of the authors and CD & PMD will serve as guarantors for the contents of this paper. CD is supported by a grant from the UK Medical Research Council (MRC) which forms part of the MRC Integrative Epidemiology Unit (IEU) at the University of Bristol (MC_UU_00032/06). DR and PMD are supported by the Baily Thomas Charitable Fund [TRUST/VC/AC/SG/6075-9277]. PMD also acknowledges support from the MRC Integrative Epidemiology Unit (IEU) at the University of Bristol (MC_UU_00032/02)). GMK acknowledges funding support from the MRC (MC_UU_00032/06; MR/W014416/1; MR/S037675/1; and MR/Z50354X/1), the Wellcome Trust (201486/Z/16/Z and 201486/B/16/Z), and the UK National Institute of Health and Care Research (NIHR) Bristol Biomedical Research Centre (BRC) at the University Hospitals Bristol and Weston NHS Foundation Trust and the University of Bristol (NIHR 203315). HJJ, DR and PMD also acknowledges funding from the NIHR Bristol BRC (NIHR 203315). The views expressed are those of the authors and not necessarily those of the NIHR or the Department of Health and Social Care. J.F.G.U. is supported by aGW4-CAT Clinical Doctoral Fellowship (22284921) and an HEIW Welsh Clinical Academic Training Fellowship. LH is supported by an Early Career Fellowship from the Elizabeth Blackwell Institute (University of Bristol) & Rosetrees Trust.

