## Supplementary Material for "Psychotic experiences and disorders in adolescents and young adults with intellectual disabilities: Evidence from a population-based birth cohort in the United Kingdom"

### Supplementary Note 1. Information on the variables used as auxiliaries in the multiple imputation analyses.

In the present analyses we had complete records on ID (exposure) and the psychotic disorder diagnoses (outcome). The total number of individuals with complete records on exposure and psychotic disorder diagnoses was 9,407.

On this basis, for the analyses investigating the associations between ID and psychotic disorder diagnoses, we used auxiliary variables to impute the covariates. All covariates were binary variables and therefore were imputed using the logit command in STATA. Details on the auxiliary variables used can be found below.

(1) Maternal marital status: assessed via questionnaire during 8 weeks of gestation, binary measure. The measure was available in 8,491 mothers of the eligible sample (12% missingness).

(2) Home ownership status: assessed via questionnaire during 8 weeks of gestation, binary measure. The measure was available in 8,445 mothers of the eligible sample (13% missingness).

(3) Car ownership status: assessed via questionnaire during 8 weeks of gestation, binary measure. The measure was available in 8,458 mothers (13% missingness).

In the case of the analyses investigating the associations between ID and psychotic experiences, the total number of individuals with complete records on exposure and outcome was 4,300. We therefore imputed the covariates using the auxiliary variables outlined above and the outcome using as an auxiliary variable the psychotic disorder diagnosis. All variables were binary and were imputed using the logit command in STATA. In the case of the analyses investigating the associations between ID and the measure reflecting the persistence and frequency of psychotic experiences (0: "Not present", 1: "Low-frequency"- experiences occurring less than weekly, 2: "High-frequency"- experiences occurring weekly or daily), we followed the same process for imputation with the exception of using the mlogit command in STATA to impute the nominal outcome variable.

### Supplementary Table 1. Characteristics of individuals with and without ID in the study sample.

| | Total Sample | Sample without ID[1] | Sample with ID[1] |
|---|---|---|---|
| Variable: | N=9,407 | N=9,071 | N=336 |
| Affective psychosis diagnosis based on GP records | 32 (0.3%) | A[2] | ≤5 |
| Psychotic experiences until 18 and/or 24 not attributed to sleep or fever | | | |
| Not present | 3,721 (39.6%) | 3,579 (39.5%) | 142 (42.3%) |
| Present | 579 (6.2%) | 555 (6.1%) | 24 (7.1%) |
| Missing | 5,107 (54.3%) | 4,937 (54.4%) | 170 (50.6%) |
| Psychotic experiences until 18 and/or 24 not attributed to sleep or fever, distressing or frequent | | | |
| Not present | 4,003 (42.6%) | 3,851 (42.5%) | 152 (45.2%) |
| Present | 297 (3.2%) | 283 (3.1%) | 14 (4.2%) |
| Missing | 5,107 (54.3%) | 4,937 (54.4%) | 170 (50.6%) |
| Trajectory of psychotic experiences | | | |
| No psychotic experiences | 1,555 (16.5%) | A[2] | A[2] |
| Persistent High | 22 (0.2%) | A[2] | ≤5 |
| Persistent Low | 135 (1.4%) | A[2] | ≤5 |
| Transient | 414 (4.4%) | A[2] | A[2] |
| Missing | 7,281 (77.4%) | A[2] | A[2] |
| sex | | | |
| male | 4,665 (49.6%) | 4,479 (49.4%) | 186 (55.4%) |
| female | 4,732 (50.3%) | 4,582 (50.5%) | 150 (44.6%) |
| Missing | 10 (0.1%) | 10 (0.1%) | 0 (0.0%) |
| parity | | | |
| <=1 | 6,511 (69.2%) | 6,290 (69.3%) | 221 (65.8%) |
| >=2 | 1,677 (17.8%) | 1,601 (17.6%) | 76 (22.6%) |
| Missing | 1,219 (13.0%) | 1,180 (13.0%) | 39 (11.6%) |
| Maternal education | | | |
| Not University graduate | 6,988 (74.3%) | 6,705 (73.9%) | 283 (84.2%) |
| University graduate | 940 (10.0%) | 931 (10.3%) | 9 (2.7%) |
| Missing | 1,479 (15.7%) | 1,435 (15.8%) | 44 (13.1%) |
| Maternal age at delivery, mean (SD) | 28.0 (4.9) | 28.0 (5.0) | 28.0 (4.9) |
| Major financial problems | | | |
| Not present | 6,280 (66.8%) | 6,055 (66.8%) | 225 (67.0%) |
| Present | 1,063 (11.3%) | 1,021 (11.3%) | 42 (12.5%) |
| Missing | 2,064 (21.9%) | 1,995 (22.0%) | 69 (20.5%) |
| Maternal anxiety during pregnancy, mean (SD) | 4.9 (3.5) | 4.9 (3.5) | 5.2 (3.6) |
| Maternal depression during pregnancy, EPDS≥12 | | | |
| Not present | 6,305 (67.0%) | 6,098 (67.2%) | 207 (61.6%) |
| Present | 1,334 (14.2%) | 1,259 (13.9%) | 75 (22.3%) |
| Missing | 1,768 (18.8%) | 1,714 (18.9%) | 54 (16.1%) |
| Home ownership status | | | |
| Owned | 6,201 (65.9%) | 5,991 (66.0%) | 210 (62.5%) |
| Rented | 2,034 (21.6%) | 1,944 (21.4%) | 90 (26.8%) |
| Missing | 1,172 (12.5%) | 1,136 (12.5%) | 36 (10.7%) |

| | Total Sample | Sample without ID[1] | Sample with ID[1] |
|---|---|---|---|
| **Variable**: | N=9,407 | N=9,071 | N=336 |
| Maternal marital status | | | |
| Married | 6,264 (66.6%) | 6,039 (66.6%) | 225 (67.0%) |
| Separated | 2,015 (21.4%) | 1,940 (21.4%) | 75 (22.3%) |
| Missing | 1,128 (12.0%) | 1,092 (12.0%) | 36 (10.7%) |
| Traumatic experiences between ages 5-11 | | | |
| Not present | 5,044 (53.6%) | 4,872 (53.7%) | 172 (51.2%) |
| Present | 2,475 (26.3%) | 2,357 (26.0%) | 118 (35.1%) |
| Missing | 1,888 (20.1%) | 1,842 (20.3%) | 46 (13.7%) |
| 1: Intellectual Disabilities<br>2: the value cannot be presented to avoid secondary disclosure. | | | |

### Supplementary Table 2a. Characteristics of the sample with complete records on exposure, psychotic disorder diagnoses and covariates.

| | Total Sample[1] | Sample with complete records[2] | Sample with incomplete records[3] |
|---|---|---|---|
| | N=9,407 | N=6,245 | N=3,162 |
| Probable ID[2] indicated by two or more sources (IQ<85) | | | |
| Not present | 9,071 (96.4%) | 6,010 (96.2%) | 3,061 (96.8%) |
| Present | 336 (3.6%) | 235 (3.8%) | 101 (3.2%) |
| Affective psychosis diagnosis based on GP records | | | |
| Not present | 9,375 (99.7%) | 6,231 (99.8%) | 3,144 (99.4%) |
| Present | 32 (0.3%) | 14 (0.2%) | 18 (0.6%) |
| Psychotic experiences until 18 and/or 24 not attributed to sleep or fever | | | |
| Not present | 3,721 (39.6%) | 2,850 (45.6%) | 871 (27.5%) |
| Present | 579 (6.2%) | 412 (6.6%) | 167 (5.3%) |
| Missing | 5,107 (54.3%) | 2,983 (47.8%) | 2,124 (67.2%) |
| Psychotic experiences until 18 and/or 24 not attributed to sleep or fever, distressing or frequent | | | |
| Not present | 4,003 (42.6%) | 3,057 (49.0%) | 946 (29.9%) |
| Present | 297 (3.2%) | 205 (3.3%) | 92 (2.9%) |
| Missing | 5,107 (54.3%) | 2,983 (47.8%) | 2,124 (67.2%) |
| Trajectory of psychotic experiences | | | |
| No psychotic experiences | 1,555 (16.5%) | 1,266 (20.3%) | 289 (9.1%) |
| Persistent High | 22 (0.2%) | 13 (0.2%) | 9 (0.3%) |
| Persistent Low | 135 (1.4%) | 107 (1.7%) | 28 (0.9%) |
| Transient | 414 (4.4%) | 328 (5.3%) | 86 (2.7%) |
| Missing | 7,281 (77.4%) | 4,531 (72.6%) | 2,750 (87.0%) |
| sex | | | |
| male | 4,665 (49.6%) | 3,132 (50.2%) | 1,533 (48.5%) |
| female | 4,732 (50.3%) | 3,113 (49.8%) | 1,619 (51.2%) |
| Missing | 10 (0.1%) | 0 (0.0%) | 10 (0.3%) |
| parity | | | |
| <=1 | 6,511 (69.2%) | 5,037 (80.7%) | 1,474 (46.6%) |
| >=2 | 1,677 (17.8%) | 1,208 (19.3%) | 469 (14.8%) |
| Missing | 1,219 (13.0%) | 0 (0.0%) | 1,219 (38.6%) |
| Maternal education | | | |
| Not University graduate | 6,988 (74.3%) | 5,445 (87.2%) | 1,543 (48.8%) |
| University graduate | 940 (10.0%) | 800 (12.8%) | 140 (4.4%) |
| Missing | 1,479 (15.7%) | 0 (0.0%) | 1,479 (46.8%) |
| Maternal age at delivery, mean (SD) | 28.0 (4.9) | 28.6 (4.7) | 26.5 (5.3) |
| Major financial problems | | | |
| Not present | 6,280 (66.8%) | 5,370 (86.0%) | 910 (28.8%) |
| Present | 1,063 (11.3%) | 875 (14.0%) | 188 (5.9%) |
| Missing | 2,064 (21.9%) | 0 (0.0%) | 2,064 (65.3%) |
| Maternal anxiety during pregnancy, mean (SD) | 4.9 (3.5) | 4.8 (3.5) | 5.6 (3.7) |
| Maternal depression during pregnancy, EPDS≥12 | | | |

| | | | |
|---|---|---|---|
| Not present | 6,305 (67.0%) | 5,261 (84.2%) | 1,044 (33.0%) |
| Present | 1,334 (14.2%) | 984 (15.8%) | 350 (11.1%) |
| Missing | 1,768 (18.8%) | 0 (0.0%) | 1,768 (55.9%) |
| Home ownership status | | | |
| Owned | 6,201 (65.9%) | 4,982 (79.8%) | 1,219 (38.6%) |
| Rented | 2,034 (21.6%) | 1,176 (18.8%) | 858 (27.1%) |
| Missing | 1,172 (12.5%) | 87 (1.4%) | 1,085 (34.3%) |
| Maternal marital status | | | |
| Married | 6,264 (66.6%) | 4,941 (79.1%) | 1,323 (41.8%) |
| Separated | 2,015 (21.4%) | 1,230 (19.7%) | 785 (24.8%) |
| Missing | 1,128 (12.0%) | 74 (1.2%) | 1,054 (33.3%) |
| Traumatic experiences between ages 5-11 | | | |
| Not present | 5,044 (53.6%) | 3,924 (62.8%) | 1,120 (35.4%) |
| Present | 2,475 (26.3%) | 1,966 (31.5%) | 509 (16.1%) |
| Missing | 1,888 (20.1%) | 355 (5.7%) | 1,533 (48.5%) |

1: Sample with data on exposure and at least one outcome measure (psychotic disorder or psychotic experiences)
2: Sample with complete records on exposure, psychotic disorder, and covariates
3: Sample with incomplete records on exposure, psychotic disorder, and covariates

### Supplementary Table 2b. Characteristics of the sample with complete records on exposure, psychotic experiences, and covariates.

| | Total Sample[1] | Sample with complete records[2] | Sample with incomplete records[3] |
|---|---|---|---|
| | N=9,407 | N=3,262 | N=6,145 |
| Probable ID indicated by two or more sources (IQ<85) | | | |
| Not present | 9,071 (96.4%) | 3,132 (96.0%) | 5,939 (96.6%) |
| Present | 336 (3.6%) | 130 (4.0%) | 206 (3.4%) |
| Affective psychosis diagnosis based on GP records | | | |
| Not present | 9,375 (99.7%) | 3,253 (99.7%) | 6,122 (99.6%) |
| Present | 32 (0.3%) | 9 (0.3%) | 23 (0.4%) |
| Psychotic experiences until 18 and/or 24 not attributed to sleep or fever | | | |
| Not present | 3,721 (39.6%) | 2,850 (87.4%) | 871 (14.2%) |
| Present | 579 (6.2%) | 412 (12.6%) | 167 (2.7%) |
| Missing | 5,107 (54.3%) | 0 (0.0%) | 5,107 (83.1%) |
| Psychotic experiences until 18 and/or 24 not attributed to sleep or fever, distressing or frequent | | | |
| Not present | 4,003 (42.6%) | 3,057 (93.7%) | 946 (15.4%) |
| Present | 297 (3.2%) | 205 (6.3%) | 92 (1.5%) |
| Missing | 5,107 (54.3%) | 0 (0.0%) | 5,107 (83.1%) |
| Trajectory of psychotic experiences | | | |
| No psychotic experiences | 1,555 (16.5%) | 1,266 (38.8%) | 289 (4.7%) |
| Persistent High | 22 (0.2%) | 13 (0.4%) | 9 (0.1%) |
| Persistent Low | 135 (1.4%) | 107 (3.3%) | 28 (0.5%) |
| Transient | 414 (4.4%) | 328 (10.1%) | 86 (1.4%) |
| Missing | 7,281 (77.4%) | 1,548 (47.5%) | 5,733 (93.3%) |
| sex | | | |
| male | 4,665 (49.6%) | 1,424 (43.7%) | 3,241 (52.7%) |
| female | 4,732 (50.3%) | 1,838 (56.3%) | 2,894 (47.1%) |
| Missing | 10 (0.1%) | 0 (0.0%) | 10 (0.2%) |
| parity | | | |
| <=1 | 6,511 (69.2%) | 2,723 (83.5%) | 3,788 (61.6%) |
| >=2 | 1,677 (17.8%) | 539 (16.5%) | 1,138 (18.5%) |
| Missing | 1,219 (13.0%) | 0 (0.0%) | 1,219 (19.8%) |
| Maternal education | | | |
| Not University graduate | 6,988 (74.3%) | 2,666 (81.7%) | 4,322 (70.3%) |
| University graduate | 940 (10.0%) | 596 (18.3%) | 344 (5.6%) |

| | | | |
|---|---|---|---|
| Missing | 1,479 (15.7%) | 0 (0.0%) | 1,479 (24.1%) |
| Maternal age at delivery, mean (SD) | 28.0 (4.9) | 29.4 (4.5) | 27.1 (5.0) |
| Major financial problems | | | |
| Not present | 6,280 (66.8%) | 2,845 (87.2%) | 3,435 (55.9%) |
| Present | 1,063 (11.3%) | 417 (12.8%) | 646 (10.5%) |
| Missing | 2,064 (21.9%) | 0 (0.0%) | 2,064 (33.6%) |
| Maternal anxiety during pregnancy, mean (SD) | 4.9 (3.5) | 4.5 (3.4) | 5.2 (3.6) |
| Maternal depression during pregnancy, EPDS≥12 | | | |
| Not present | 6,305 (67.0%) | 2,823 (86.5%) | 3,482 (56.7%) |
| Present | 1,334 (14.2%) | 439 (13.5%) | 895 (14.6%) |
| Missing | 1,768 (18.8%) | 0 (0.0%) | 1,768 (28.8%) |
| Home ownership status | | | |
| Owned | 6,201 (65.9%) | 2,807 (86.1%) | 3,394 (55.2%) |
| Rented | 2,034 (21.6%) | 423 (13.0%) | 1,611 (26.2%) |
| Missing | 1,172 (12.5%) | 32 (1.0%) | 1,140 (18.6%) |
| Maternal marital status | | | |
| Married | 6,264 (66.6%) | 2,691 (82.5%) | 3,573 (58.1%) |
| Separated | 2,015 (21.4%) | 550 (16.9%) | 1,465 (23.8%) |
| Missing | 1,128 (12.0%) | 21 (0.6%) | 1,107 (18.0%) |
| Traumatic experiences between ages 5-11 | | | |
| Not present | 5,044 (53.6%) | 2,021 (62.0%) | 3,023 (49.2%) |
| Present | 2,475 (26.3%) | 1,212 (37.2%) | 1,263 (20.6%) |
| Missing | 1,888 (20.1%) | 29 (0.9%) | 1,859 (30.3%) |

1: Sample with data on exposure and at least one outcome measure (psychotic disorder or psychotic experiences)
2: Sample with complete records on exposure, psychotic experiences, covariates
3: Sample with incomplete records on exposure, psychotic experiences, covariates

### Supplementary Table 3. Associations between ID, psychotic disorders, and psychotic experiences in complete record analyses.

| Outcome | Model | N | Sample with outcome | Sample with exposure | Sample with exposure and outcome | OR[1] (95% CIs[2]) | Prevalence of outcome in sample without ID[7] | Prevalence of outcome in sample with ID[7] | Risk difference |
|---|---|---|---|---|---|---|---|---|---|
| Affective psychosis diagnosis based on GP records | Unadjusted[3] | 9407 | 32 | 336 | ≤5[4] | 3.89 (1.36, 11.16) | A[8] | A[8] | 0.88 (-0.28, 2.05) |
| Affective psychosis diagnosis based on GP records | Unadjusted[5] | 6245 | 9 | 130 | ≤5[4] | 7.05 (1.95, 25.45) | A[8] | A[8] | 1.09 (-0.35, 2.53) |
| Affective psychosis diagnosis based on GP records | Adjusted[6] | 6245 | 9 | 130 | ≤5[4] | 9.23 (2.44, 34.96) | A[8] | A[8] | 1.45 (-0.43, 3.33) |
| Psychotic experiences until 18 and/or 24 not attributed to sleep or fever | Unadjusted[3] | 4300 | 579 | 166 | 24 | 1.09 (0.70, 1.70) | 13.43 (12.43, 14.51) | 14.46 (9.99, 20.93) | 1.03 (-4.42, 6.48) |
| Psychotic experiences until 18 and/or 24 not attributed to sleep or fever | Unadjusted[5] | 3262 | 412 | 130 | 19 | 1.19 (0.73, 1.96) | 12.55 (11.44, 13.76) | 14.62 (9.65, 22.14) | 2.07 (-4.11, 8.25) |
| Psychotic experiences until 18 and/or 24 not attributed to sleep or fever | Adjusted[6] | 3262 | 412 | 130 | 19 | 1.17 (0.70, 1.93) | 12.10 (10.99, 13.32) | 14.11 (9.28, 21.46) | 1.76 (-4.33, 7.85) |
| Psychotic experiences until 18 and/or 24 not attributed to sleep or fever, distressing or frequent | Unadjusted[3] | 4300 | 297 | 166 | 14 | 1.25 (0.72, 2.20) | 6.85 (6.12, 7.66) | 8.43 (5.11, 13.92) | 1.59 (-2.71, 5.88) |
| Psychotic experiences until 18 and/or 24 not attributed to sleep or fever, distressing or frequent | Unadjusted[5] | 3262 | 205 | 130 | 10 | 1.26 (0.65, 2.43) | 6.23 (5.43, 7.13) | 7.69 (4.24, 13.95) | 1.47 (-3.19, 6.12) |
| Psychotic experiences until 18 and/or 24 not attributed to sleep or fever, distressing or frequent | Adjusted[6] | 3262 | 205 | 130 | 10 | 1.24 (0.64, 2.42) | 5.92 (5.13, 6.82) | 7.33 (4.02, 13.35) | 1.37 (-3.25, 6.00) |

1: Odds Ratio
2: Confidence Interval
3: Complete records on exposure and outcome
4: the value cannot be presented to avoid secondary disclosure.
5: Complete records on exposure, outcome, covariates
6: Adjusted for: child sex (male/female), maternal parity (≤1 child versus ≥ 2 children), major financial problems in the family when the child was 8 months old (yes/no), maternal highest educational attainment (32 weeks gestation), maternal age (at delivery), maternal Crown-Crisp anxiety scores (18 weeks gestation), maternal depression measured with the Edinburgh Postnatal Depression Scale (EPDS; 18 weeks gestation scores ≥ 13).
7: Intellectual Disability
8: Exact value omitted to avoid disclosure of cell counts <5 (3.8 = 100 5/130, where 130 is the number with ID in complete record analyses)

Supplementary Table 4. Associations between ID and the four longitudinal profiles of psychotic experiences in complete record analyses.

| Trajectory | model | N | RRR[1] (95% CIs[2]) |
|---|---|---|---|
| No psychotic experiences | Unadjusted[3] | 2126 | Ref |
| Persistent High | | 2126 | 3.28 (0.74, 14.45) |
| Persistent Low | | 2126 | 1.26 (0.49, 3.23) |
| Transient | | 2126 | 1.58 (0.91, 2.72) |
| No psychotic experiences | Unadjusted[4] | 1714 | Ref |
| Persistent High | | 1714 | 2.93 (0.37, 23.17) |
| Persistent Low | | 1714 | 0.67 (0.16, 2.82) |
| Transient | | 1714 | 1.45 (0.76, 2.78) |
| No psychotic experiences | Adjusted[5] | 1714 | Ref |
| Persistent High | | 1714 | 2.32 (0.28, 19.18) |
| Persistent Low | | 1714 | 0.69 (0.16, 2.95) |
| Transient | | 1714 | 1.39 (0.72, 2.68) |

1: Relative Risk Ratio
2: Confidence Interval
3: Sample with complete records on exposure, outcome
4: Sample with complete records on exposure, outcome, covariates.
5: Adjusted for: child sex (male/female), maternal parity (≤1 child versus ≥ 2 children), major financial problems in the family when the child was 8 months old (yes/no), maternal highest educational attainment (32 weeks gestation), maternal age (at delivery), maternal Crown-Crisp anxiety scores (18 weeks gestation), maternal depression measured with the Edinburgh Postnatal Depression Scale (EPDS; 18 weeks gestation scores ≥ 13).

Supplementary Table 5. Complete records mediation analyses with childhood trauma for the associations between ID, psychotic disorders, and psychotic experiences.

| outcome | Model | N | TCE[1]; OR[2] (95%CIs[3]) | NDE[4]; OR[2] (95%CIs[3]) | NIE[5]; OR[2] (95%CIs[3]) |
|---|---|---|---|---|---|
| Affective psychosis diagnosis based on GP records | Unadjusted[6] | 7519 | 5.95 (1.83, 19.28) | 5.44 (1.69, 17.47) | 1.09 (0.99, 1.20) |
| Affective psychosis diagnosis based on GP records | Unadjusted[7] | 5890 | 6.79 (1.83, 25.25) | 6.11 (1.64, 22.75) | 1.11 (0.96, 1.28) |
| Affective psychosis diagnosis based on GP records | Adjusted[8] | 5890 | 8.97 (2.35, 34.29) | 8.20 (2.17, 30.91) | 1.09 (0.96, 1.25) |
| Psychotic experiences until 18 and/or 24 not attributed to sleep or fever | Unadjusted[6] | 7519 | 1.09 (0.68, 1.75) | 1.06 (0.67, 1.69) | 1.03 (0.98, 1.08) |
| Psychotic experiences until 18 and/or 24 not attributed to sleep or fever | Unadjusted[7] | 5890 | 1.19 (0.72, 1.97) | 1.16 (0.71, 1.90) | 1.03 (0.98, 1.09) |
| Psychotic experiences until 18 and/or 24 not attributed to sleep or fever | Adjusted[8] | 5890 | 1.16 (0.71, 1.89) | 1.14 (0.70, 1.85) | 1.02 (0.98, 1.07) |
| Psychotic experiences until 18 and/or 24 not attributed to sleep or fever, distressing or frequent | Unadjusted[6] | 7519 | 1.30 (0.70, 2.41) | 1.26 (0.68, 2.32) | 1.03 (0.97, 1.09) |
| Psychotic experiences until 18 and/or 24 not attributed to sleep or fever, distressing or frequent | Unadjusted[7] | 5890 | 1.26 (0.61, 2.60) | 1.21 (0.59, 2.49) | 1.04 (0.97, 1.11) |
| Psychotic experiences until 18 and/or 24 not attributed to sleep or fever, distressing or frequent | Adjusted[8] | 5890 | 1.24 (0.60, 2.55) | 1.21 (0.59, 2.47) | 1.03 (0.97, 1.09) |

1: Total effect
2: Odds Ratio
3: Confidence Intervals
4: Natural Direct Effect
5: Natural Indirect Effect
6: Sample with complete records on exposure, mediator, outcome
7: Sample with complete records on exposure, mediator, outcome, covariates.
8: Adjusted for: child sex (male/female), maternal parity (≤1 child versus ≥ 2 children), major financial problems in the family when the child was 8 months old (yes/no), maternal highest educational attainment (32 weeks gestation), maternal age (at delivery), maternal Crown-Crisp anxiety scores (18 weeks gestation), maternal depression measured with the Edinburgh Postnatal Depression Scale (EPDS; 18 weeks gestation scores ≥ 13).
